# Molecular epidemiology of rifampicin resistant *Mycobacterium tuberculosis* in Vietnam

**DOI:** 10.64898/2026.04.20.26351312

**Authors:** Ori E. Solomon, Viet Nhung Nguyen, Thu Anh Nguyen, Hoa Binh Nguyen, Emily Lai-Ho MacLean, Greg J. Fox, Marcel A. Behr

## Abstract

**Background:** Vietnam is a top 20 burden country for multi-drug resistant/rifampicin-resistant tuberculosis (MDR/RR-TB), with nearly 10,000 cases a year. With the emergence of new diagnostic assays for *M. tuberculosis* and resistance, along with new drugs for both treatment and prevention, we sought to better understand the molecular epidemiology of RR-TB in this high-burden setting, through the study of clinical trial isolates from the VQUIN MDR trial.

**Methods:** We assembled a sample of cultured isolates, collected from patients with confirmed RR-*M. tuberculosis* within 10 provinces, enriching for isolates from outside of the 2 major cities, Hanoi and Ho Chi Minh City. We subjected these isolates whole genome sequencing (WGS) and bioinformatic analysis, with a subset subject to phenotypic drug susceptibility testing to evaluate phenotypic/genotypic concordance. New genome sequences were phylogenetically contextualised to publicly-available *M. tuberculosis* genome sequences sampled in Vietnam from National Center for Biotechnology Information (NCBI) Sequence Read Archives (SRA).

**Results:** Isolates from 252 RR-TB cases passed quality controls and were available for analysis. Xpert MTB/RIF had a high concordance with WGS-based rifampicin-resistance prediction (PPV=96.8%). Of the 244 isolates confirmed to be rifampicin resistant, a high proportion (235/244 = 96.3%) had mutations associated with resistance to at least one other first- or second-line antibiotic. Phenotypic drug susceptibility testing (DST) for rifampicin, isoniazid, and levofloxacin was completed for 77 isolates with a high concordance demonstrated between DST and genomic-based resistance predictions (67/77, 87.0% RIF; 76/77, 98.7% INH; 73/77, 94.8%LFX). High concordance was also observed with new and repurposed antibiotics linezolid (100%, 60/60), pretomanid (100%, 60/60), and bedaquiline (56/60, 93.3%). Rifampicin-resistant strains were more likely to be lineage 2.2.1, compared to rifampicin-susceptible *M. tuberculosis* strains in Vietnam, particularly in the major cities.

**Conclusions:** The high prevalence of secondary drug-resistance beyond RIF and INH, along with the dominance of one major lineage across geographic regions, provides insights on the spread of MDR/RR-TB in Vietnam and reinforces the importance of prompt and broad detection of drug-resistance to inform the timely initiation of effective drug regimens.

## Introduction

Drug-resistant tuberculosis (TB) is a leading cause of morbidity and mortality from an antibiotic resistant infection with an estimated 150,000 deaths^1^. These deaths are largely attributable to multi-drug resistant/rifampicin-resistant tuberculosis (MDR/RR-TB), which affects approximately 400,000 people each year globally^1^. The WHO has highlighted the importance of scaling up diagnostics and treatments in 20 high incidence countries of MDR/RR-TB, which make up 86% of incident cases annually^2^. Vietnam ranked eleventh globally in incidence of drug-resistant TB, with 9900 new RR-TB notifications in 2023^1^. In addition to the high RR-TB burden, Vietnam boasts a varied *Mycobacterium tuberculosis* ecology, where 3 main endemic lineages (lineages 1, 2, and 4) are represented. Given that different lineages have been reported to have a different propensity to develop resistance, or to be directly associated with resistance-associated mutations, the analysis of different lineages provides a window into the evolution of drug resistance.

Vietnam has widely implemented rapid molecular assays such as Xpert MTB/RIF and Xpert MTB/RIF Ultra (hitherto referred to as Xpert) for simultaneous screening of bacteriological evidence of TB disease and rifampicin (RIF) resistance^3^. In 2024, 94% of new cases and 100% of previously-treated cases were tested for RIF susceptibility^4^. Based on this testing, 4.4% of patients with new bacteriologically confirmed pulmonary TB and 16.8% of patients with previously-treated pulmonary TB had RR-TB^4^. Because the most recent national level anti-tuberculosis drug resistance survey from 2011 restricted to first-line antibiotics^5^, estimates of the rate of antibiotic resistance to other antibiotics across Vietnam are largely unknown. In 2024, Nguyen et al. conducted a descriptive analysis of RR*-M. tuberculosis* in Vietnam, combining whole genome sequencing (WGS) and phenotypic data to screen for resistance, and finding that resistance was mainly confined to first-line antibiotics^6^. This study provided valuable insights, however, it was limited to patients from the two largest cities, Hanoi and Ho Chi Minh City^5^, where only a third of annual RR-TB cases reside, and it did not test phenotypic susceptibility to new and repurposed antibiotics. As new diagnostic technologies (WGS and targeted next generation sequencing) are introduced and new antibiotics for the treatment of MDR/RR-TB are scaled up, data on the epidemiology of resistance and concordance of diagnostic modalities is crucial for effective care.

In the years 2016-2021, a placebo-controlled clinical trial (VQUIN) for the safety and efficacy of levofloxacin (LFX)-based TB preventive treatment in contacts of MDR/RR-TB cases was conducted in Vietnam. Consecutively diagnosed patients with MDR/RR-TB from 10 provinces, including its two largest cities and large rural centers, were invited to enroll in the study^7^. We used whole genome sequencing data from isolates derived from these patients to identify resistance to first-line and second-line antibiotics, including new and repurposed drugs. We used phenotypic drug susceptibility testing (pDST) to quantify concordance between phenotypic and genotypic assays on a subsample of this cohort, highly enriched for exposure to drugs used in the treatment of MDR-TB. We also characterized the phylogeographical distribution of RR-TB across the country to elucidate patterns in the emergence of second-line antibiotic resistance.

## Methods

### Sampling

Methods used to recruit participants in 10 provinces of Vietnam have been described^8^. In patients with RR-TB detected by Xpert screening, expectorated sputum samples were collected for microbiological study (index isolates). Additionally, household contacts who were recruited to the trial were followed up for 24 months post-randomization. Additional samples were obtained from household contacts who either had microbiologically confirmed TB at screening (co-prevalent TB) or developed microbiologically confirmed TB within the 30 months after enrolment (incident TB). Isolates were obtained following N-acetyl-L-cysteine-sodium hydroxide decontamination and Mycobacterial Growth Indicator Tube broth culture, then frozen at −80°C for subsequent genomic and phenotypic analysis at the Research Institute of the McGill University Health Centre (Montrel, Canada). We implemented a quasi-randomized subsampling strategy. Firstly, available index case isolates were randomly selected with a geographical distribution proportional to recruitment in the VQUIN trial (n=200). For provinces where random selection resulted in less than 10 isolates, we enriched the sampling to achieve a minimum of 10 per province (n=32), except for Quang Nam Province that only had 2 available isolates. In addition to this sampling of index isolates from across the country, we included all available isolates from the contacts of patients with TB who developed TB (n=39) and their respective index case isolate, where available (n=32 pairs). Patient demographics were obtained from the parent trial.^7,9^

### Bacterial culture and whole genome sequencing

Isolates were re-grown in Lowenstein-Jensen Media or Middlebrook 7H10 agar media supplemented with 0.5% glycerol and 10% oleic acid-albumin-dextrose-catalase. Genomic DNA was extracted using the QIAamp UCP Pathogen lysis kit (Qiagen, Hildon, Germany) with a modified mechanical lysis protocol, as described previously^10^. Paired-end sequencing libraries were prepared using the S4 reagent kit and shotgun sequencing performed using Novaseq 6000 with read lengths of 150bp and a mean number of reads of 22M reads/sample (range 6.4M-70.6M).

### Phenotypic testing

Phenotypic testing of a sub-sample of isolates was performed using the agar proportion method, as described previously^11^. Three key drugs in determining standard treatment recommendations were tested using the WHO revised recommended critical concentrations for RIF (0.5 ug/mL), high-level isoniazid (INH) resistance (1.0 ug/mL), and LFX (1.0 ug/mL).

Susceptibility to new and repurposed drugs bedaquiline (BDQ), pretomanid (PMD), and linezolid (LZD) was tested using the resazurin-based microtiter assay. Isolates were passaged in Middlebrook 7H9 media supplemented with 0.2% glycerol, 0.1% Tween-80, and 10% albumin-dextrose-catalase (7H9 complete media) and grown to mid-logarithmic phase. Cultures were then resuspended to an OD_600_ of 0.003 and dispensed (50 µL) in transparent flat-bottom 96-well plates containing twofold serial dilutions of each drug in 50 µL 7H9 complete media, as well as one column that served as a media control and a drug free control. Final antibiotic concentrations upon inoculation with bacteria ranged from 1.9×10^-4^ µg/mL – 100 µg/mL. Plates were incubated for 6 days at 37°C. Resazurin was added (0.025% w/v in each well) and plates incubated overnight. Fluorescence was measured with excitation and emission at 560mm and 590mm, respectively, using a Tecan Infinite M Plex to quantify metabolism of resazurin to resorufin as indicator of cell viability. MIC was determined using the Gompertz equation in GraphPad Prism (v10). Isolates were called resistant at MIC shifts of ≥4x MIC of H37Rv. All isolates were tested in biological and technical duplicates.

### Bioinformatic analysis

#### Read filtering and decontamination

FastQC^12^ was used to assess sequence data quality. Kraken was used to taxonomically classify reads with a custom database comprising of bacteria and the human genome (GRCh38). Reads classified as *Mycobacterium tuberculosis complex* (TAXID: 77643) were extracted using KrakenTools^13^. Sequencing runs with >10% mycobacterial reads that were not classified as *Mycobacterium tuberculosis complex* at the genus level were deemed contaminated and filtered out. Raw paired-end reads were quality trimmed with Trimmomatic v0.40 to remove low-quality reads.

#### Variant calling, in-silico resistance and lineage calling, and phylogenetics

Variant calling was performed using Snippy v4.6.0^14^ and sequences were aligned to the *M. tuberculosis* H37Rv reference genome (GenBank: NC_000962.3) with default parameters. Isolates with coverage <90% and depth of coverage <100x were excluded. A whole genome multiple sequence alignment was generated using snippy-core and processed use snippy-clean_full_aln, with polymorphic sites identified using snp-sites^15^. Regions of low confidence and PE/PPE genes were masked.

A SNV based maximum-likelihood phylogenetic tree construction was performed using IQ-TREE v2.3.6^16^. Tree inference was done using the general time reversible model with gamma-distributed rate heterogeneity and ascertainment bias correction to account for use of variable sites only, with 1000 ultrafast bootstrap replicates for branch support. Genomic based antibiotic resistance prediction and lineage annotation was performed using TB-profiler v6.6.2^17^. The phylogenetic tree was annotated with the interactive tree of life (https://itol.embl.de/). To enrich our database, we searched for all available genomes from NCBI SRA for *M. tuberculosis* sequences samples in Vietnam. Sequences were downloaded from bioprojects: PRJNA355614, PRJNA1028637, PRJDB8553, PRJDB8544, PRJDB10923, PRJNA1044389^18–22^. Sequences were processed and filtered as mentioned above. Mixed infections were excluded from phylogenetic tree analysis. Lineage 3 isolates were also excluded from phylogenetic tree as lineage 3 is rare in Vietnam and not believed to be endemic to the country.

A pairwise-comparison matrix across all isolates was done using Snp-dists v0.8.3 (https://github.com/tseemann/snp-dists), excluding low-confidence regions^23^ and antibiotic resistance associated genes. Using alignments of our available isolates, we used a strict cutoff of 5 single nucleotide polymorphisms (SNPs) and a loose cutoff of 12 SNPs to either support or rule out transmission to household contacts.

### Statistical analysis

To evaluate geographical differences in lineage distribution of MDR/RR-*M. tuberculosis*, isolates were grouped into 2 groups: the major cities (Hanoi and Ho Chi Minh City) and the rest of the country. Relative risk was calculated for the risk of sublineage 2.2.1 infection compared to any other sublineage in these cities compared to the rest of the country, significance was tested using Fisher’s test, and p-value <0.05 was determined significantly different.

## Results

### Study population

The sample set assembled had a total of 303 isolates, of which 287 (94.7%) were successfully sequenced and passed quality control and mycobacterial read decontaminations. These 287 comprised 252 index isolates and 35 contact isolates. Geographic distribution and patient demographics regarding these are provided in Table 1. Index cases were older and more likely to be male than their household contacts. A majority of index cases (69.4%) had reported a previous episode of TB (Table 1).

**Table 1.** Demographic and clinical characteristics of sequenced isolates.

| Characteristic | Index cases<br>(N=252) | Household contacts<br>(N=35) |
| --- | --- | --- |
| **Province** |  |  |
| **Northern region** |  |  |
| Hanoi | 68 (27.0%) | 11 (31.4%) |
| Nam Dinh | 14 (5.5%) | 0 (0%) |
| Thanh Hoa | 14 (5.5%) | 2 (5.7%) |
| **Central region** |  |  |
| Da Nang | 15 (6.0%) | 1 (2.9%) |
| Khanh Hoa | 15 (6.0%) | 1 (2.9%) |
| Quảng Nam | 1 (0.4%) | 0 (0%) |
| **Southern region** |  |  |
| An Giang | 14 (5.5%) | 4 (11.4%) |
| Can Tho | 11 (4.4%) | 1 (2.9%) |
| Ho Chi Minh City | 84 (33.3%) | 13 (37.1%) |
| Tien Giang | 16 (6.3%) | 2 (5.7%) |
| **Age at baseline** |  |  |
| Mean (SD) | 45.0 (15.5) | 38.6 (14.2) |
| Median (IQR) | 44.4(33.2, 57.4) | 35.6 (30.8, 46.7) |
| **Participant gender** |  |  |
| Male | 185 (73.4%) | 18 (51.4%) |
| Female | 67 (26.6%) | 17 (48.6%) |
| **Baseline HIV status** |  |  |
| Negative | 238 (94.4%) | 35 (100.0%) |
| Positive | 14 (5.6%) | 0 (0.0%) |
| **Baseline smear status** |  |  |
| Smear negative | 58 (23.0%) | NA |
| Smear positive | 175 (69.4%) | NA |
| Unavailable | 19 (7.5%) | NA |
| **Comorbidities** |  |  |
| None | 143 (56.7%) | 24 (68.6%) |
| Chronic kidney disease | 3 (1.2%) | 0 (0.0%) |
| Chronic lung disease | 4 (1.6%) | 1 (2.9%) |
| Diabetes | 51 (20.2%) | 2 (5.7%) |
| Liver disease | 14 (5.6%) | 0 (0.0%) |
| Other condition | 25 (9.9%) | 8 (22.9%) |
| Missing | 12 (4.8%) | 0 (0.0%) |
| **History of TB** |  |  |
| No prior episodes | 77 (30.6%) | 32 (91.4%) |
| At least one previous episode | 175 (69.4%) | 3 (8.6%) |

NA: Not applicable

### Comparison of molecular resistance prediction and phenotypic drug susceptibility testing

We first set out to determine inter-assay concordance in three modalities of drug-resistance detection. Although the inclusion criteria for index cases required a RIF-resistance report by molecular testing (primarily Xpert), we found 8 index case strains did not have any mutations in the RIF resistance determining region (RRDR) detected by WGS and therefore had been classified as false positives (Xpert PPV compared to WGS: 96.8%; Table 3). One isolate had a RIF associated mutation in *rpoB* resulting in an amino acid substitution outside of the RRDR (V170P) that was falsely captured by Xpert. Using RIF resistance as a proxy for the presence of INH resistance, we found that the PPV of Xpert for MDR-TB – that is, resistant to both RIF and INH - was 91.9%.

Of the 252 index cases, we used the proportion method to test for phenotypic resistance to RIF, INH, and LFX in 77 isolates from 9 districts/provinces that had undergone WGS-based genotyping (Table 2).

**Table 2:** Phenotypic-genotypic resistance concordance of 77 isolates to RIF, INH, and LFX. S: susceptible, R: resistant.

| **RIF** | **Phenotypic S** | **Phenotypic R** | **Total** |
| --- | --- | --- | --- |
| **Genotypic S** | 4 | 0 | 4 |
| **Genotypic R** | 10 | 63 | 73 |
| **Total** | 14 | 63 | 77 |
| **INH** |  |  |  |
| **Genotypic S** | 8 | 1 | 9 |
| **Genotypic R** | 0 | 68 | 68 |
| **Total** | 8 | 69 | 77 |
| **LFX** |  |  |  |
| **Genotypic S** | 57 | 1 | 58 |
| **Genotypic R** | 3 | 16 | 19 |
| **Total** | 60 | 17 | 77 |

For RIF, the phenotype-genotype concordance was 67/77 (87.0%). The 4 isolates randomly selected that had no RRDR mutation (false-positive Xpert) were confirmed to be phenotypically susceptible. Of the 10 discordant results, 7 were due to disputed RRDR mutations (D435Y, H445C, H445G, and L430P). Two additional strains had RRDR mutations with uncertain significance: a double mutation in M434L and D435G and a double amino acid in-frame deletion at Q426_P427.

For INH, the phenotype-genotype concordance was 76/77 (98.7%). All isolates with a canonical INH-R mutation had a resistant phenotype, while 8/9 isolates with no identified INH-R mutation were susceptible. In the single discordant case of INH susceptibility, we detected a mutation in *katG* (G299S) of uncertain significance.

For LFX, the phenotype-genotype concordance was 73/77 (94.8%). 17 isolates showed concordant resistance. Of the 4 discordant isolates, 3 phenotypically susceptible isolates had resistance-associated mutations in *gyrA* (A90V and D94A) whereas one phenotypically resistant isolate had no resistance associated mutations in *gyrA* or *gyrB*.

Taken together, the results of phenotypic testing of 77 isolates for the 3 antibiotics showed an overall concordance of 93.5% (216/231) with WGS. Of the 15 discordant results, 7 were associated with disputed mutations – known to cause discordance – for which current guidelines^24^ recommend the priority to the genotypic result and 4 had mutations of uncertain significance.

### pDST and Putative resistance mutations to new and repurposed antibiotics

A random subsample of sequenced isolates also underwent pDST for new and repurposed antibiotics BDQ, PMD, and LZD by REMA (n=60). We found a perfect concordance of phenotypic susceptibility and genotypic resistance prediction for PMD and LZD, with all isolates showing comparable or lower MICs than H37Rv (0.17 ug/mL and 1.0 ug/mL, respectively). BDQ had a lower accuracy, with 56/60 (91.8%) of isolates having a susceptible phenotype and no known resistance-associated mutations. In one of the four isolates with an elevated MIC compared to H37Rv (≥4x MIC of H37Rv; 0.10 ug/mL), a non-synonymous polymorphism was identified in the BDQ-resistance associated gene *Rv0678* (D8A), not previously described in the WHO catalogue. Putative resistance associated mutations in other isolates with a BDQ resistance phenotype were not identified. One isolate with an elevated BDQ MIC was on a pre-XDR *M. tuberuclosis* genomic background (resistant to RIF, INH, and fluroquinolones), fulfilling the definition of extensively drug resistant (XDR) phenotype.

### Prevalence and distribution of antibiotic-resistant genotypes across Vietnam

Having validated our results with the antibiotics above, we explored resistance prediction to 10 additional antibiotics, in both the index patients (n=252) and their contacts (n=35) (Table 3).

**Table 3:** Genotypic resistance prediction in Index patients and household contacts. RIF= rifampicin, INH= isoniazid, EMB= ethambutol, PZA=pyrazinamide, LFX=levofloxacin, BDQ= bedaquiline, PMD= pretomanid, LZD= linezolid, STR= streptomycin, AMI= amikacin, KAN= kanamycin, CAP= capreomycin, CFZ= clofazimine, ETH= ethionamide, PAS= Para-aminosalicylic acid, CSR= cycloserine. HHC= Household contact

| Antibiotic | Index (%) | HHC (%) | Total (%) |
| --- | --- | --- | --- |
| **RIF** | 244 (96.8) | 14 (40) | 258 (89.9) |
| **INH** | 232 (92.1) | 21 (60) | 253 (88.2) |
| **EMB** | 170 (67.5) | 10 (28.6) | 180 (62.7) |
| **PZA** | 147 (58.3) | 8 (22.9) | 155 (54) |
| **LFX** | 49 (19.4) | 4 (11.4) | 53 (18.5) |
| **BDQ** | 3 (1.2) | 0 | 3 (1) |
| **PMD** | 0 | 0 | 0 |
| **LZD** | 0 | 0 | 0 |
| **STR** | 221 (87.7) | 22 (62.9) | 243 (84.7) |
| **AMI** | 8 (3.2) | 0 | 8 (2.8) |
| **KAN** | 11 (4.4) | 0 | 11 (3.8) |
| **CAP** | 6 (2.4) | 0 | 6 (2.1) |
| **CFZ** | 3 (1.2) | 0 | 3 (1) |
| **ETH** | 36 (14.3) | 5 (14.3) | 41 (14.3) |
| **PAS** | 8 (3.2) | 0 | 8 (2.8) |
| **CSR** | 1 (0.4) | 0 | 1 (0.3) |
| **Resistance profile** |  |  |  |
| **Susceptible** | 4 (1.6) | 11 (31.4) | 15 (5.2) |
| **HR-TB** | 3 (1.2) | 8 (22.9) | 11 (3.8) |
| **RR-TB** | 11 (4.4) | 1 (2.9) | 12 (4.2) |
| **MDR-TB** | 184 (73.0) | 9 (25.7) | 193 (67.2) |
| **Pre-XDR-TB** | 49 (19.4) | 4 (11.4) | 53 (18.5) |
| **Other** | 1 (0.4) | 2 (5.7) | 3 (1.0) |

Among index RR-TB isolates, we observed a high frequency of resistance to first-line antibiotics: INH (92.1%), ethambutol (EMB, 67.5%), pyrazinamide (PZA, 58.3%) and streptomycin (STR, 87.7%). RIF resistance without INH resistance was a rare occurrence (n=15; 6%); of these 15, 5 were otherwise pan-susceptible, 5 were STR-resistant, 1 was EMB-resistant and 4 were LFX-resistant. The most common genotype for first-line antibiotic susceptibility was resistance to all 5 first-line antibiotics (Fig1A). Furthermore, in RR-TB index cases, we observed 29.9% resistance to at least one second-line antibiotic, including 19.4% LFX, 14.3% ethionamide (ETH), 3.2% amikacin (AMK), 4.4% kanamycin (KAN), 2.4% capreomycin (CAP), and 1.2% clofazimine (CFZ) and 1.2% bedaquiline (BDQ). Patterns of drug-resistance were mapped geographically by district (Fig 1B) demonstrating nationwide presence of LFX resistance and the presence of pre-XDR-TB strains in the north, central and southern regions.

**Figure 1.**
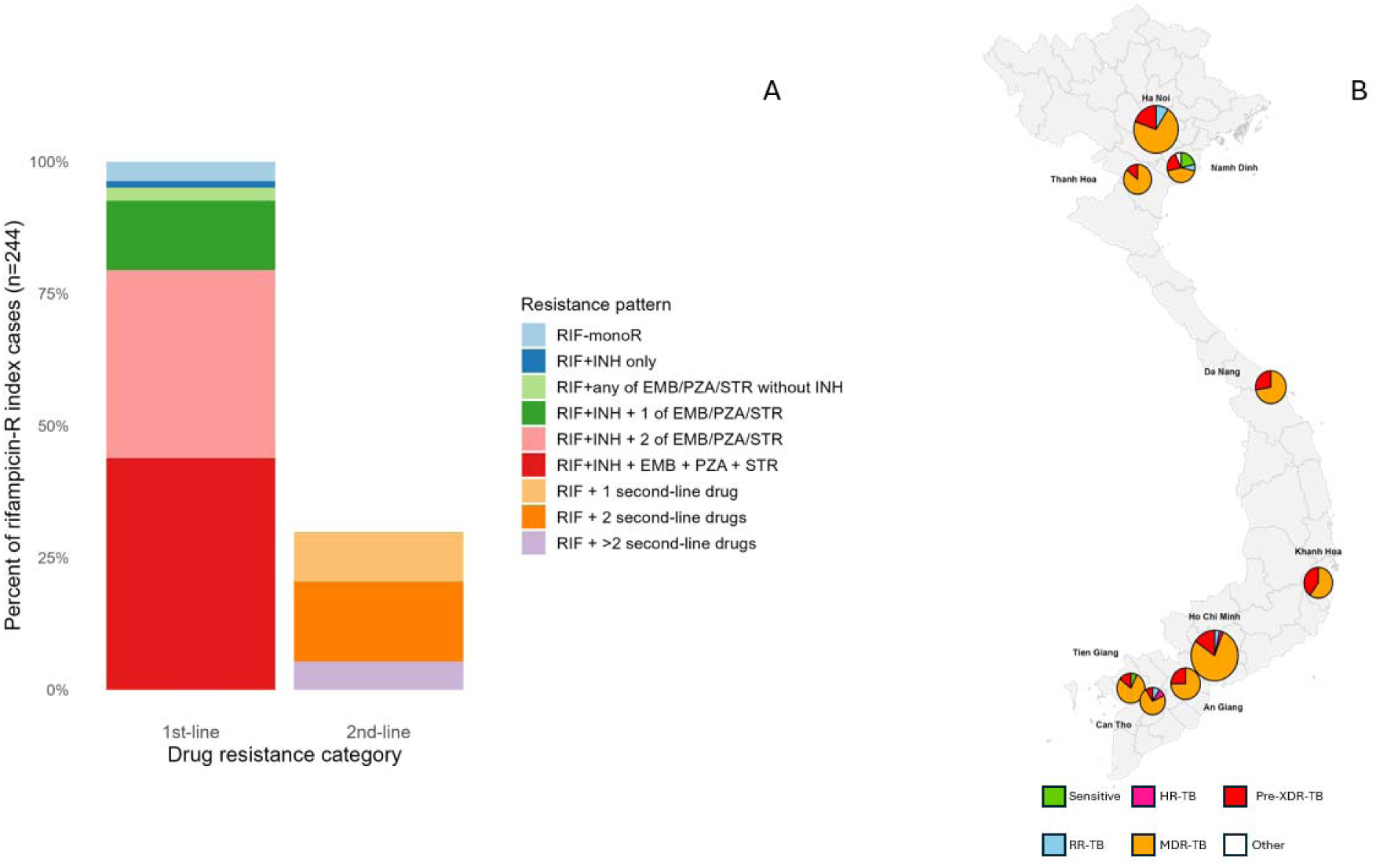
Genotypic prediction of antibiotic resistance profiles amongst 252 index cases from the VQUIN MDR trial. A) Patterns of resistance to first- and second-line antibiotics amongst RR-TB index cases. B) Proportion of resistance profiles in each district/region sampled. Resistance categories labeled according to the WHO definitions. Quang Nam not shown as it had a single representative isolate (MDR-TB).

Amongst isolates collected from household contacts (n=35), the frequency of resistance to first-line antibiotics was lower: INH (60%), RIF (40%) EMB (28.5%), PZA (23%) and STR (63%). Of household isolates, monomono23% were INH monoresistant, 38% MDR-TB, 11.7% pre-XDR-TB, and a single case was RIF monoresistant.

Heteroresistance (co-occurrence of susceptible and resistant populations within a host) to any antibiotic was detected in 26 index isolates and 1 household isolate (27/289, 9.3%). The most frequent antibiotics associated with heteroresistance were RIF, EMB, PZA, and LFX (n=10, 7, 8, 12, respectively). However, the antibiotics with the highest proportion of resistance strains harbouring heteroresistance were LFX and BDQ (12/43 and 2/3, respectively). We also looked at how many strains had two or more mixed allele loci (2 or more unfixed SNPs at a frequency <90% in the same antibiotic-resistance associated gene) to a specific antibiotic as a marker of ongoing selection for resistance, with RIF (4/10), EMB (2/7), STR (2/2), and LFX (2/12) resistance genes being the most represented. Heteroresistance was more commonly associated with minority alleles (below 50%) than majority alleles (>=50%). No heteroresistance to AMK, PAS, CAP, or cycloserine (CSR) was detected.

### Transmission of drug-resistant M. tuberculosis in the household

Combining genotypic and epidemiological data, we set out to identify clustering of drug-resistant TB. We successfully sequenced 28 household contacts of 25 unique index cases, for a total of 30 (out of 32 available) pairwise household contact-index comparisons. In combining all isolates (index and household contacts) and using a strict cutoff (5SNPs), we identified 22 clusters made up of two isolates and 1 cluster made up of three isolates, with a median distance of 2 SNPs. In the same total cohort and using the loose cutoff (12 SNPS), we identified 18 additional clusters, including an additional 49 isolates, in which transmission could not be ruled out. At this higher cutoff, there were 41 total clusters, including 31 clusters of two isolates and 10 clusters ranging from 3-6 isolates, with a median distance of 9 SNPs. Of the household contact strains, 17 clustered in the strict cutoff analysis and 19 clustered in the soft cutoff analysis. When looking at for clustering of index-household pairs, only 12 household contacts’ strains (42.8%) were found in clusters that also contained an additional member of their household in the strict cutoff, with 2 additional household containing clusters that could not be ruled out with the loose cutoff (Fig 2).

**Figure 2.**
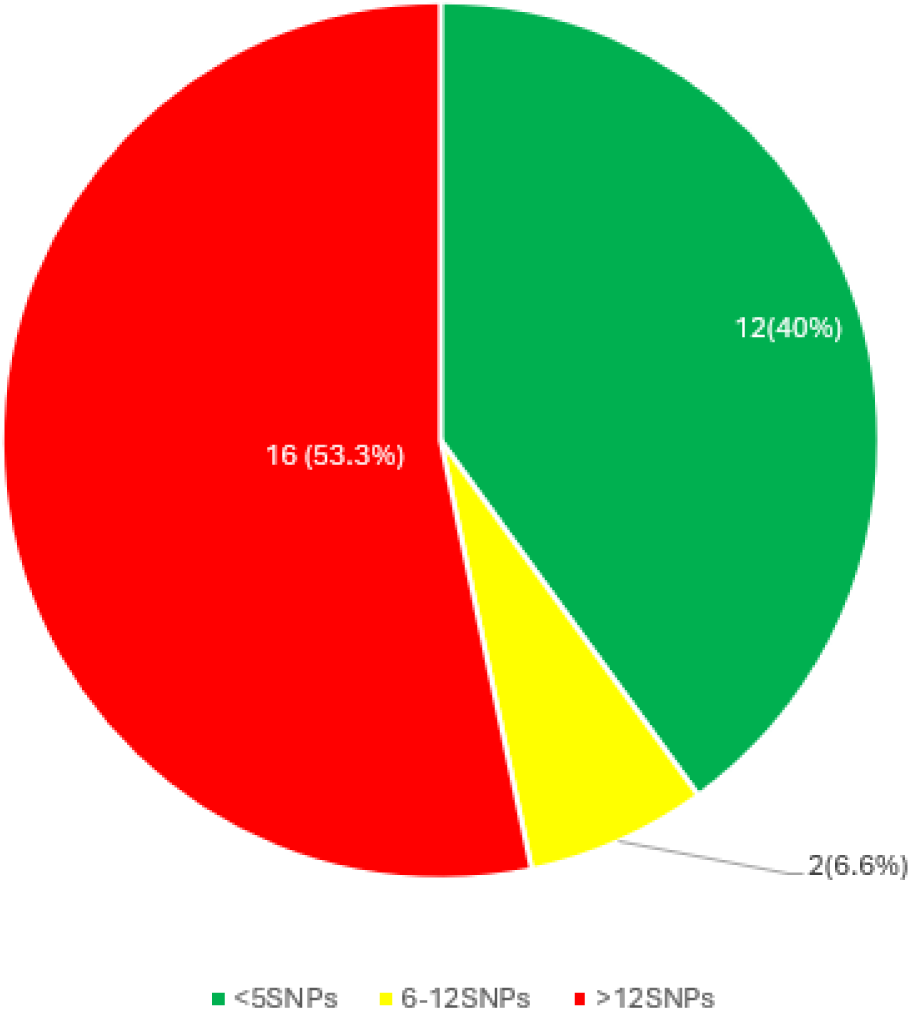
Clusters containing household contacts. Sequenced isolate pairs with known epidemiological link (household contacts; n=30). Red: >12 SNPs between pair, ruling out household transmission. Yellow: 6-12SNPs loose cutoff for potential household transmission. Green: <6SNPs strict cutoff for potential household transmission.

### Phylogenetics of antibiotic-resistant *M. tuberculosis*

We were able to sequence isolates from all 10 provinces that participated in the VQUIN trial. Hanoi and Ho Chi Minh City were the most represented regions, with 77 and 97 isolates represented, respectively (Table 1).

A phylogenetic tree was constructed from the 252 index isolates to better understand the genetic diversity of MDR/RR-TB isolates and main lineages were mapped across 9 of the participating regions/provinces (excluding the single isolate available from Quang Nam Province) (Fig. 3). A combination of lineages 1,2 and 4 were detected in northern, central, and southern regions of the country, with all 3 detected in most populous provinces (Fig3A). Lineage 2 was the dominant lineage of RR/MDR-*Mtb* in every province, making up 88.5% of all strains sequenced.

**Figure 3:**
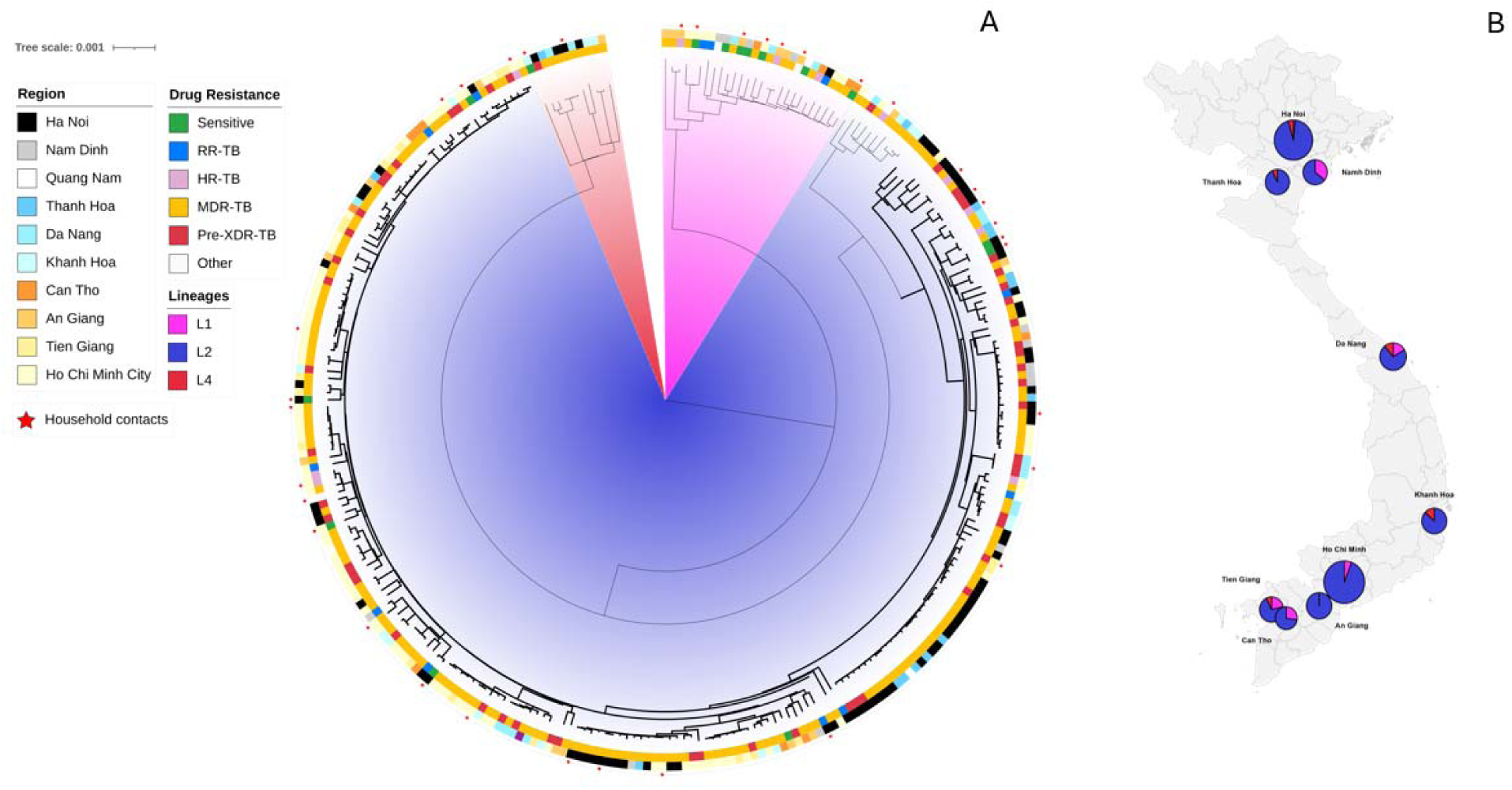
Geography of main *M. tuberculosis* lineages in Vietnam. A) Maximum-liklihood tree of 287 *M. tuberculosis* isolates from the VQUIN MDR trial. Nodes in bold indicate lineage 2.2.1. Red stars used to indicate isolates from household contacts B) Proportion of lineages represented in each region sampled. Quang Nam was excluded as it only had a single representative isolate (sublineage 2.2.1).

**Figure 4.**
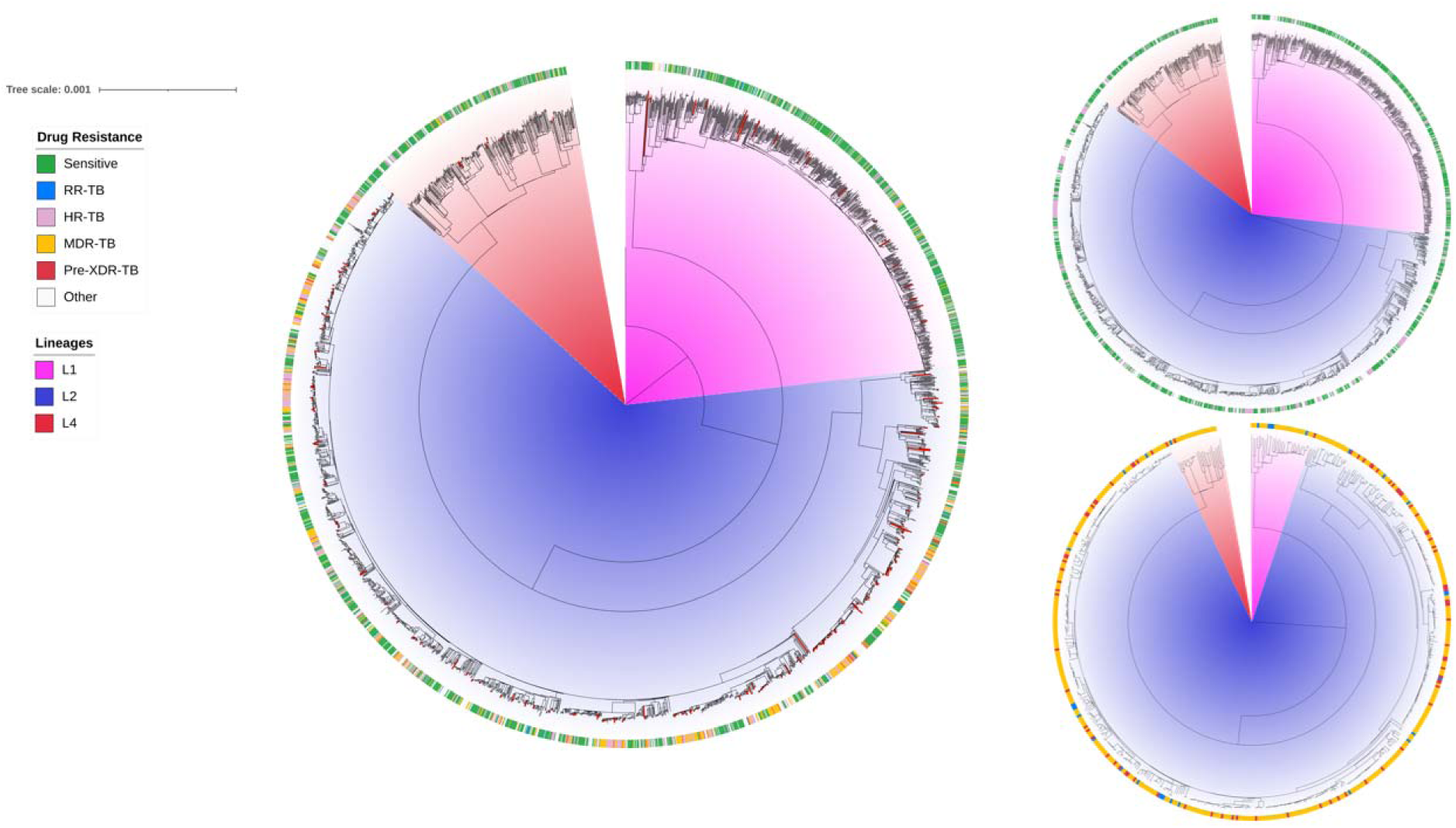
Phylogenies of enriched Vietnam dataset to contextualize newly sequenced isolates. Left: Phylogeny of 3351 *M. tuberculosis* strains across Vietnam sampled from 2003-2022. Terminal branches in red mark isolates from VQUIN trial. Top right: Phylogeny of RIF susceptible strains (n=2787). Bottom Right: Phylogeny of RIF resistant strains (n=564).

Sublineage 2.2.1 made up 92% of all Lineage 2 isolates, with few sublineage 2.1 and other 2.2 sublineages detected. MDR/RR-TB was more likely to be due to a lineage 2.2.1 infection in the major cities (Hanoi and Ho Chi Minh City) than in the rest of the country (RR 1.2, 95% CI 1.1-1.4; p-value = 0.0041).

To contextualize our RR-TB cohort within the greater Vietnam *M. tuberculosis* ecological context, we enriched our Vietnam isolate dataset with 3062 isolates previously sampled in Vietnam from 2004-2022, primarily from Ho Chi Minh City, with additional sequences available from Hanoi and Da Nang. While lineage 2 remained the dominant lineage, prevalence dropped from 88.5% in the VQUIN trial to 65.4% when also including drug sensitive TB. When dividing all obtained isolates from Vietnam by RIF-resistance, lineage 2 represented 90.3% of RIF-R strains and 60.3% of RIF-S strains. LFX resistance was even more strongly associated with lineage 2 in Vietnam, with 81 pre-XDR-TB strains from lineage 2, a single isolate from lineage 4, and no lineage 1 isolates found to have LFX resistance.

## Discussion

Our analysis of nearly 300 RR-*M. tuberculosis* strains from Vietnam has reinforced the importance of resistance to many drugs, including INH, PZA, EMB, STR, and fluoroquinolones. Through WGS, we were able to show that many case-contact pairs did not have matching genomes, indicating that community circulation is driving much of the spread of these strains. The finding that resistance is observed across geographic regions, in different lineages and sublineages, supports the notion that resistance is emerging and re-emerging de novo, with a selection towards this happening most commonly in lineage 2 strains, which are encountered across the country. Maintaining efficacy of new and repurposed drugs, such as a BDQ, relies on robust phenotypic susceptibility testing to detect resistance in strains with non-canonical resistance mutations, particularly in patients who would benefit from second-line antibiotic regimens.

Although next generation sequencing-based resistance prediction is being rolled out globally, its effective implementation relies on integration into the current diagnostic landscape that is complementary to existing technologies used in resistance detection. The two modalities of antibiotic susceptibility testing recommended by the WHO are molecular assays, such as the GeneXpert platform, and phenotypic testing using the proportion method or commercial broth systems. As expected, a majority of RR-TB isolates had mutations in the RRDR, corresponding to a high concordance between WGS and Xpert. However, TB-profiler called 2 strains as resistant despite a lack of phenotypic evidence found in the literature. Mutations outside of the RRDR were also detected, including compensatory mutations and non-RRDR disputed mutations such as RpoB_I491F and RpoB_V170P. Conflictingly, one strain was incorrectly included in the analysis by Xpert as there were no mutations identified in the RRDR, however it was genotypically predicted to be resistant due to the V170P mutation in RpoB. False-positive calls by Xpert have been reported in similar screenings in Vietnam^6^.

Phenotypic-genotypic concordance was undertaken for six antimycobacterials. These antibiotics were selected as current WHO guidelines for the treatment of pan-susceptible TB and MDR-TB heavily rely on susceptibility to these antibiotics. We found the strongest genotype-phenotype concordance in PMD (100%), LZD (100%) and INH (98.7%), followed by LFX (94.8%), BDQ (91.8%) and RIF (87.0%). RIF-resistance discordance was most commonly attributed to disputed mutations in *rpoB.* These mutations, found within and external to the RRDR, have been associated with low-level resistance to RIF that is near the critical concentration, frequently generating a susceptible pDST result, although showing an association with higher rates of treatment failure^25–30^. These disputed mutations were found in 17.5% of index isolates across the country. Three additional discordances in RIF resistance were due to three separate mutations with unknown significance. Although we screened for high-INH resistance, there were no isolates with suspected low-level resistance detected, indicated by a resistance associated genotype and a susceptible phenotype. Once case of INH-resistance discordance was observed, putatively linked to a SNP in *katG* with an uncertain significance. LFX resistance discordance was observed amongst 4 isolates, 3 of which had canonical resistance-associated mutations in the *gyrA* quinolone resistance determining region (QRDR) also observed in phenotypically resistant strains and have been shown to cause higher rates of genotypic-phenotypic discordance compared to other QRDR mutations^31^. The other discordant strain produced a resistant phenotype, however, mutations associated with resistance were not observed in the QRDR of *gyrA* or *gyrB*. In the new and repurposed drugs, we did not find any resistance phenotypes to LZD and PMD. Only one isolate resistant phenotype could be linked with a putative novel resistance mutation in Rv0678. No putative resistance mutations were detected in the other resistant phenotypes, one of which having an XDR-TB phenotype, despite not being detectable by WGS.

Similarly to Nguyen et al.^6^, we found high rates of genotypic resistance to first-line antibiotics amongst RR-TB cases, consistent with other reports from Southeast Asia^32,33^. Of note, we found the rate of pre-XDR-TB amongst RR-TB patients to be at 19.4%, and 3 cases of BDQ resistant genotypes were also detected, although these were not detected in pre-XDR-TB isolates. Household contacts of MDR/RR-TB patients were at a higher risk of developing MDR/RR-TB and pre-XDR-TB compared to numbers reported for the general population at 40% and 11.4%, respectively^34^. INH monoresistance was observed in 23% of household contacts, suggesting an increase from the 10% INH monoresistance rate reported among all isolates tested in the 2015 national survey^5^. Heteroresistance was most frequently found to RIF, EMB, PZA, and LFX, with LFX and BDQ resistance being most commonly found in heterogeneous populations, an indication of ongoing selective pressure and resistance evolution across first- and second-line antibiotics, including new classes such as BDQ.

Using a cutoff of 12 SNPs, we ruled out transmission of MDR/RR-TB to 53.3% of household contact cases (Fig2), suggesting more than half of drug-resistant infections are acquired outside of the household of the recognised index case. This finding is similar to rates of household transmission reported in the VQUIN trial^7^ as isolates used in the trial were also included in our analysis, with the dataset expanded to include all available household contact strains.

Lineages 1, 2, and 4 are known to be widely circulating in Vietnam, with all three lineages found in the northern, central, and southern regions. Lineage 2 is known to be the most common lineage found in Vietnam, and in this MDR/RR-TB enriched cohort was the predominant lineage. Using publicly available *M. tuberculosis* genomes available on NCBI SRA, isolates from the VQUIN trial were plotted to alongside >3000 genomes sampled from Vietnam between 2003-2022, a time range that spans over the duration of the trial. Whereas lineage 2 made up 65.4% of all isolates, it made up 90.3% of RR-TB strains. As was seen in the RIF resistant VQUIN cohort, lineage distribution is not uniform across all regions, and thus oversampling from large urban centres may bias our findings. This combined with regional differences observed within our cohort underscore the importance of conducting regular molecular epidemiology surveillance at regional/district level to better inform local public health interventions.

In conclusion, in a study of RR-TB isolates from a clinical trial undertaken across Vietnam, a concordance of WGS-based resistance prediction with Xpert and phenotypic testing to six antibiotics was noted. This evidence can inform the national TB program in its ongoing roll out of next generation sequencing-based antibiotic resistance prediction technology. Furthermore, household transmission was ruled out for a half of contacts, confirming that community transmission is a key driver in the ongoing RR-TB epidemic. This emphasises the importance of expanding case detection beyond targeted high-risk groups such as household contacts in order to interrupt community transmission^35^. Finally, we observed changes in phylogenetic structure of RR-TB strains, with a high enrichment for lineage 2 and an underrepresentation of lineages 1 and 4 in RR-TB. Understanding the epidemiology of drug-resistant *M. tuberculosis* in high-burden settings such as Vietnam is crucial to developing locally-appropriate case-detection strategies and halting transmission of TB in the community.

## Data Availability

All data produced in the present study are available upon reasonable request to the authors

## Acknowledgements

We acknowledge the collective efforts of the staff at 10 Provincial and City Hospitals across the Vietnam National TB Program network participating in this study. This project was funded by a National Health and Medical Research Council (NHMRC) Project Grant (1081443) to GJF and a Foundation Grant from the Canadian Institutes for Health research (CIHR) to MAB (FDN 148362). GJF was supported by a NHMRC Leadership Fellowship (2007920). ELM was supported by a CIHR Fellowship (472823). MAB was supported by a Tier 1 Canada Research Chair.

